# Use of recently vaccinated individuals to detect bias in test-negative case-control studies of COVID-19 vaccine effectiveness

**DOI:** 10.1101/2021.06.23.21259415

**Authors:** Matt D.T. Hitchings, Joseph A. Lewnard, Natalie E. Dean, Albert I. Ko, Otavio T. Ranzani, Jason R. Andrews, Derek A.T. Cummings

## Abstract

Post-authorization observational studies play a key role in understanding COVID-19 vaccine effectiveness following the demonstration of efficacy in clinical trials. While bias due to confounding, selection bias, and misclassification can be mitigated through careful study design, unmeasured confounding is likely to remain in these observational studies. Phase III trials of COVID-19 vaccines have shown that protection from vaccination does not occur immediately, meaning that COVID-19 risk should be similar in recently vaccinated and unvaccinated individuals, in the absence of confounding or other bias. Several studies have used the estimated effectiveness among recently vaccinated individuals as a negative control exposure to detect bias in vaccine effectiveness estimates. In this paper we introduce a theoretical framework to describe the interpretation of such a bias-indicator in test-negative studies, and outline assumptions that would allow the use of recently vaccinated individuals to correct bias due to unmeasured confounding.

## Introduction

The test-negative case-control study (TNCC) is a common observational study design for estimating vaccine effectiveness, including vaccines against influenza^1,2^, rotavirus^3,4^, and other infectious diseases^5^. With worldwide roll-out of vaccines against COVID-19 in progress, TNCCs have been a key tool for assessing the direct effect of vaccination on individuals in real-world settings, within groups not well represented in clinical trials, effectiveness against outcomes other than primary trial outcomes, and effectiveness against variants of concern.

Valid estimates of vaccine effectiveness are obtained from TNCCs when (i) vaccination has no effect on incidence of the test-negative condition, (ii) misclassification of disease etiology is minimized, and (iii) bias from other sources is minimized^6,7^. Previous work^8,9^ has quantified the bias arising from differences in exposure, susceptibility, and healthcare-seeking between unvaccinated and vaccinated populations (manifesting as confounding and/or selection bias), misclassification of test-positive or test-negative individuals, and differential buildup of naturally-acquired immunity over time among the vaccinated and unvaccinated. Such bias is likely to be exacerbated in many situations in which COVID-19 vaccines are being evaluated, with priority for vaccine given to individuals at highest risk of severe disease, individual utilization of vaccine being associated with perceptions of risk or prior exposure^10–12^, and high spatiotemporal heterogeneity in vaccine coverage, infection risk, and access to testing^13–15^.

Test-negative controls are an example of a negative control outcome^16,17^ used to reduce bias in observational studies, but additional negative control outcomes or exposures can uncover remaining biases. A test-negative study of influenza vaccination in seniors used hospitalization before and after the influenza season as negative control outcomes to detect bias in influenza vaccine effectiveness estimates^18^. Ideally, a negative control outcome used as a “bias-indicator” is an outcome that shares as many common causes as possible with the pathogen of interest, except for vaccination. An association between vaccination and the negative control outcome suggests differences in disease risk between vaccinated and unvaccinated individuals that is unrelated to vaccine effectiveness. Under various assumptions, negative control outcomes have been used to reduce or correct bias^19,20^. A negative control exposure such as vaccination for an unrelated pathogen, not causing the outcome but sharing unmeasured confounders with the exposure of interest, can be used in a similar way^21,22^.

As immune response to COVID-19 vaccines takes time to develop, and individuals can be infected before vaccination but develop symptoms within the incubation period, protection is not expected to manifest immediately following vaccination^23,24^. Therefore, for observational studies of COVID-19 vaccines, recent vaccination has been used as a negative control exposure to detect bias^25–29^. Here we describe how time-variant differences between vaccinated and unvaccinated individuals, as well as changes in risk over time within vaccinated individuals, would manifest among recently vaccinated individuals and discuss the utility of this group as a bias-indicator. In addition, we outline the assumptions necessary for using recent vaccination to correct for bias in vaccine effectiveness in later time periods.

## Methods

### Theoretical framework

We follow the framework from Lewnard et al^8^ for a test-negative case-control study. Specifically, we assume that a clinical syndrome of interest (e.g., acute respiratory illness, ARI) could arise from SARS-CoV-2 infection (+) or from infection from causes unrelated to SARS-CoV-2 (-). Individuals undergo constant hazard *λ*_*i*_ of infection, where subscript *i* denotes the test-positive or test-negative outcome (+ or -), and develop ARI upon infection with probability *π*_*i*_. Upon developing ARI, individuals seek treatment with probability *μ*_v_, where subscript *v* represents vaccination status. To allow for differential risk of infection among vaccinated and unvaccinated individuals independent of vaccination, we define the parameter *α*_*vi*_ to be the hazard ratio for infection among this group relative to the general population.

We extend the framework to model a vaccination campaign, in which a proportion *v* of a population of size *P* are vaccinated (V), with the remaining population unvaccinated (U). For simplicity, we consider a single-dose vaccine which provides no protection within *T*_*P*_ days of vaccination, and has efficacy given by *VE* = *φ*(1 − *θ*) thereafter. Therefore, among those who will eventually be vaccinated, at different times individuals may have not received the vaccine (pending vaccination, P), recently vaccinated (R) or fully vaccinated (F). The timescale *t* is defined relative to the start of the vaccination campaign, *t*=0. For simplicity, we assume that all individuals are vaccinated at the same time *T*_*V*_, so that they are protected at time *T*_*V*_+*T*_*P*_. Allowing each individual to have different vaccination times produces identical results if vaccination time is not associated with SARS-CoV-2 infection risk (Supplementary Materials). Table 1 displays a list of model parameters used in this framework.

**Table 1:** Table of parameters and definitions, adapted from^8^

| Parameter | Definition |
| --- | --- |
| $\lambda_i$ | Force of infection for SARS-CoV-2 (+) or non-SARS-CoV-2 pathogens (-) |
| $\pi_i$ | Probability of ARI given infection for SARS-CoV-2 (+) or non-SARS-CoV-2 pathogens (-) |
| $\varphi$ | Proportion of individuals responding to vaccine |
| $\mu_v$ | Probability of seeking treatment given ARI, among individuals who are unvaccinated ($v=U$), pending vaccination ($v=P$), recently vaccinated ($v=R$), or fully vaccinated ($v=F$) |
| $\theta$ | Hazard ratio for infection resulting from vaccine-derived protection (among responders) |
| $\alpha_{vi}$ | Hazard ratio for infection with SARS-CoV-2 (+) or non-SARS-CoV-2 pathogens (-) (relative to population average) due to factors other than vaccine-derived protection, among individuals who are unvaccinated ($v=U$), pending vaccination ($v=P$), recently vaccinated ($v=R$), or fully vaccinated ($v=F$) |
| $T_V$ | Time of vaccination, relative to start of vaccination campaign |
| $T_P$ | Time from vaccination to full protection from vaccine |
| $P$ | Total size of population |
| $v$ | Proportion of population who are vaccinated |

For simplicity, we assume that the study is conducted in a setting in which both the prevalence of SARS-CoV-2 infection and of ARI from other causes are low, to minimize misclassification of cases due to ARI of other etiologies occurring in individuals infected with SARS-CoV-2^9^. We follow the method of Lewnard et al^8^ demonstrating that, in the presence of differential exposure and susceptibility between vaccinated and unvaccinated individuals, the odds ratio (OR) of vaccination comparing test-positive to test-negative individuals estimates the following quantity:

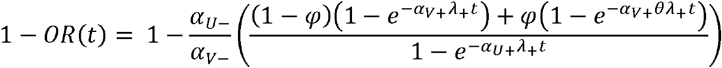

## Results

### Interpretation of the odds ratio comparing recently vaccinated to unvaccinated (“bias-indicator”)

If we assume the vaccine has no biological effect on the risk of being a case among those recently vaccinated, the cumulative incidence of the test-positive and test-negative conditions among recently vaccinated individuals arises from person-time between *T*_*V*_ and *T*_*V*_*+T*_*P*_ among those who are vaccinated.

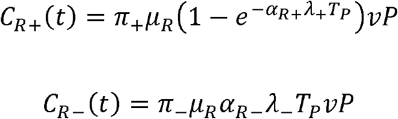

The cumulative incidence of the test-positive and test-negative conditions among unvaccinated individuals arises from person-time among those who are not vaccinated at the time of analysis, and among those who are pending vaccination. In particular,

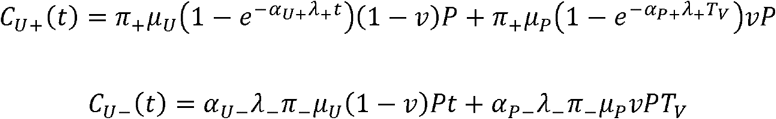

The estimated vaccine direct effect among individuals recently vaccinated, compared with those not vaccinated, could serve as an indicator of bias.

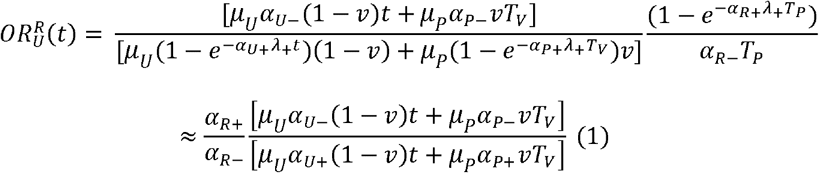

when *t* is close to *T*_*V*_, and *T*_*V*_ and *T*_*P*_ are small. Therefore, over a short time scale following initiation of the vaccine campaign, the odds ratio comparing recently vaccinated individuals to unvaccinated individuals estimates the relative susceptibility to SARS-CoV-2 infection compared to infections with another etiology among individuals eligible for and having recently received vaccination, compared to those who are not vaccinated. The bias-indicator is dependent on the proportion of individuals who are pending vaccination among unvaccinated individuals. Individuals awaiting vaccination might be more similar in their characteristics to vaccinated individuals compared to unvaccinated individuals, affecting the magnitude of the bias-indicator. In addition, the composition of the unvaccinated group might change over the course of a vaccination campaign, leading to dynamic changes in the bias-indicator.

### Interpretation of the odds ratio with recently vaccinated persons as a reference group (“bias-correction”)

Now assume that differences in exposure, susceptibility, and healthcare-seeking over time in vaccinated individuals are negligible (i.e *α*_*R*−_ = *α*_*F*−_ = *α*_*V*−_, *α*_*R*+_ = *α*_*F* +_ = *α*_*V*+_, and *μ*_*R*_ = *μ*_*F*_ = *μ*_*V*_). Then

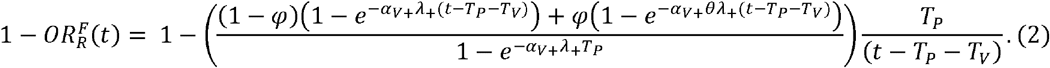

(The Supplementary Materials show a full derivation of this expression). When *t* is close to the *T*_*V*_, and *T*_*P*_ is small, equation 2 reduces to:

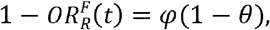

providing an unbiased estimate of vaccine efficacy.

### Illustration of bias-indicator and bias-correction method

The magnitude and direction of the bias-indicator (equation 1) is displayed in Figure 1 (left column), and the effect of applying the bias-correction method (equation 2) to the vaccine effectiveness estimate (right column). In the left column, the black line represents the bias-indicator estimated over time since vaccination. In the right column, the blue line represents the VE estimate in the presence of bias, and the red line represents the bias-corrected VE estimate. The efficacy of the vaccine is 70%.

**Figure 1.**
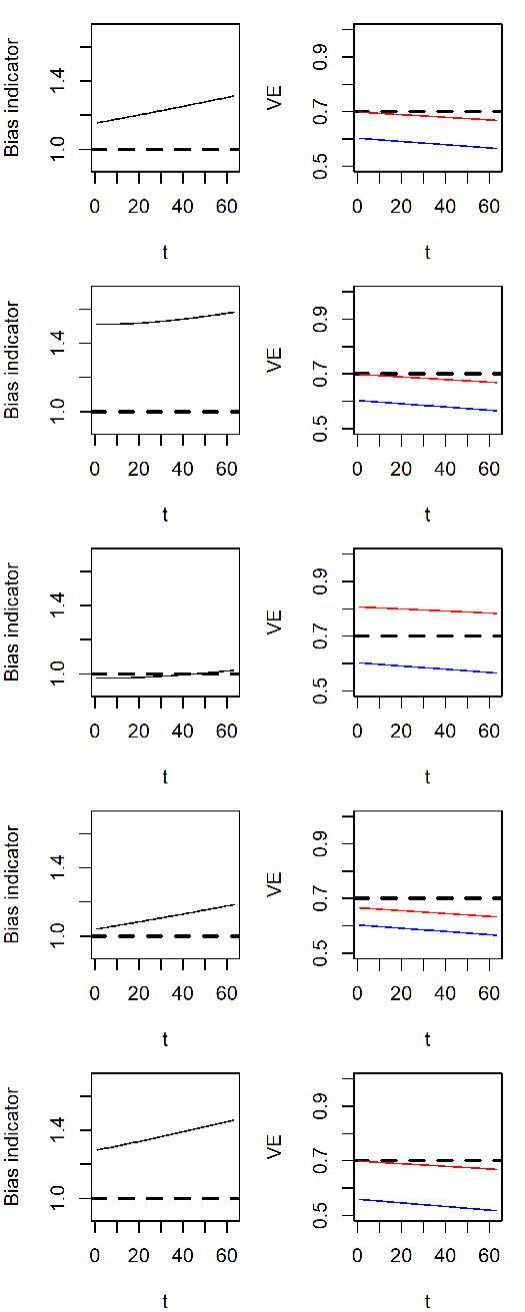
Bias-indicators (left column) and biased (blue) and bias-corrected (red) estimates of vaccine efficacy (right column) over time since vaccination. We assume higher risk of SARS-CoV-2 among vaccinated individuals that is time-invariant (first row), lower risk of SARS-CoV-2 in individuals awaiting SARS-CoV-2 vaccination (second row), higher risk of SARS-CoV-2 among fully vaccinated individuals (third row), a small effect of vaccination on disease risk among recently vaccinated individuals (fourth row), and reduced probability of seeking testing among recently and fully vaccinated individuals (fifth row).

In the first row, the relative hazard of SARS-CoV-2 comparing vaccinated to unvaccinated, from factors unrelated to vaccination, is *α*_*P+*_ *= α*_*R+*_ *=α*_*F+*_=1.25 (i.e. vaccinated individuals have higher infection risk due to factors other than vaccination). In this case, the bias-indicator is above one, the uncorrected method underestimates VE, and the bias-corrected method returns a valid estimate over the time scale considered. In the second row, the relative hazard of SARS-CoV-2 is lower among those who are pending vaccination, so that *α*_*P+*_ =0.8. Individuals that are known to have received a vaccine are unlikely to have tested positive in the weeks preceding vaccination, otherwise they would have been less likely to get vaccinated. This represents a form of immortal time bias. In this case, the magnitude of the bias-indicator is increased, while the vaccine effectiveness estimates remain the same. In the third row, both individuals pending vaccination and recently vaccinated have lower risk than those fully vaccinated and unvaccinated (representing relaxation of other risk mitigation practices by those who are fully immunized). In this case, counteracting biases cause the bias-indicator to be close to one, but the bias-correction method overestimates vaccine effectiveness because recently vaccinated individuals are at lower risk than unvaccinated individuals. In the fourth, the vaccine efficacy is 10% among recently vaccinated individuals. In this case, the bias-indicator is slightly above one, and the bias-corrected method underestimates vaccine effectiveness as the reference period includes time in which individuals were protected by vaccination. Finally, in the bottom row, recently and fully vaccinated individuals are less likely to seek care for moderate symptoms. In this case, the bias-indicator is above one, the uncorrected method underestimates vaccine effectiveness (as severe cases are overrepresented among vaccinated individuals), and the bias-correction method returns a valid estimate of vaccine effectiveness over the timescale shown. Code to reproduce these figures is provided in the Supplementary Material.

## Discussion

Comparison of recently vaccinated and unvaccinated individuals in a test-negative case-control study can be interpreted, under certain assumptions and over a short time scale following initiation of the vaccination campaign, as the relative difference in infection risk between vaccinated and unvaccinated groups due to factors other than vaccination. Furthermore, by using recently vaccinated individuals as the reference group, such a study can remove bias caused by differences in SARS-CoV-2 infection risk and susceptibility to symptoms between individuals who get vaccinated and those who do not. This method, when it is valid, could be particularly useful for studies based on secondary data sources or conducted in low-resource settings in which detailed confounder information is not available.

The interpretation of the bias-indicator and the validity of the bias-correction method rely on two key assumptions: that all parameters relating to differences in exposure, susceptibility, and test-seeking between fully vaccinated and recently vaccinated individuals are time-invariant; and that the definition of recently vaccinated is chosen such that vaccination has not had time to affect the risk of infection or the chosen clinical outcome. One can imagine situations in which the first assumption does not hold. For example, individuals who have a scheduled vaccine appointment may take extra measures to protect themselves from risk in the time immediately before or following vaccination, and conversely take fewer precautions once they believe they are protected by vaccination (α_F-_>α_R-_ and α_F+_>α_R+_). In addition, some observational studies^26,28^ have observed “deferral bias”, in which individuals feeling sick choose not to get vaccinated and subsequently test positive for SARS-CoV-2, leading to apparent protection from vaccination among recently vaccinated individuals (α_PI_>α_RI_). Finally, COVID-19 vaccination campaigns have been conducted so that high-risk individuals are initially targeted or are early adopters^27^. In such situations, there is more potential for differences between those who vaccinated earlier and later. The likelihood of being fully vaccinated among cases and controls may therefore be confounded by predictors of early vaccination. Even if controlling for risk group through matching or stratification, those who receive the vaccine earlier within eligibility groups may have different risk of disease.

For studies not restricted to severe symptoms, difference in the distribution of moderate and severe disease, and differences in test-seeking behavior, between fully vaccinated and recently vaccinated individuals, can lead to further bias if not accounted for^30^. Individuals who have been recently vaccinated may be less likely to seek testing for moderate symptoms, believing them to be side effects of vaccination. On the other hand, a fully vaccinated individual might be less likely to seek testing for moderate symptoms once they believe themselves to be protected against SARS-CoV-2 infection. Therefore, the bias-indicator may represent a mixture of time-invariant and time-varying bias that could be in either direction, and consequently the proposed bias-correction method is not guaranteed to reduce bias.

In addition, it is not clear for every COVID-19 vaccine what time period should define a recently vaccinated individual who has yet to experience clinical protection. Consistent with results from Phase III trials of COVID-19 vaccines, vaccine protection is not immediate. Differences in COVID-19 risk were observed starting 10-12 days following first dose in the trials of the BNT162b2 mRNA^23^ and the mRNA-1273 vaccines^24^, and 28 days following first dose in the interim analysis of the ChAdOx1 vaccine^31^. A natural choice would be some quantile of the incubation period of SARS-CoV-2 (e.g. 11.5 days^32^), as infections seen in this period would likely have been acquired before vaccination. If the clinical syndrome of interest is hospitalization, a longer time window would be appropriate, representing the time from infection to hospitalization. However, in populations with moderate seroprevalence, individuals who have had prior infection may experience protection from a single dose of vaccine earlier than expected based on trial results^33^. In addition, although this has not been demonstrated for any COVID-19 vaccine, some vaccines are known to elicit non-specific immune responses^34^, which could lead to some vaccine effect in the days immediately following vaccination and an underestimate of vaccine effectiveness among fully vaccinated individuals when correcting for bias. The time window should therefore be chosen to be as short as possible to minimize the possibility of bias. However, a shorter window leads to lower prevalence of “recent vaccination”, increasing the standard error of 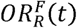. Selection of the time window for definition of “recently vaccinated” thus constitutes a trade-off between bias and variance.

This discussion underscores the need for clear reasoning if choosing recently vaccinated individuals as a reference group in observational studies in an attempt to control for unmeasured confounding. While under certain assumptions the bias can be minimized, these assumptions are likely to be unverifiable from the available data. The time period used as the reference group should be specified in the protocol with explicit justification, and the increased variance of the vaccine effectiveness estimator should be accounted for when calculating necessary sample size and assessing study feasibility. Data detailing time to onset of immunogenicity with established correlates of protection would be of value to inform design of studies comparing risk in different time periods after vaccination as a bias-correction strategy.

## Data Availability

Code for replication is available from the corresponding author via https://github.com/mhitchings/tnccbias

## Notes

Conflicts of interest: J.A.L. has received grants and consulting fees from Pfizer, Inc., unrelated to this research. The remaining authors report no conflicts of interest.

Source of financial support: This work was supported by grants R01-AI14812701 from the National Institute for Allergy and Infectious Diseases to J.A.L., and R01-AI139761 from the National Institute for Allergy and Infectious Diseases to N.E.D.

### Competing Interest Statement

J.A.L. has received grants and consulting fees from Pfizer, Inc., unrelated to this research. The remaining authors report no conflicts of interest.

### Funding Statement

This work was supported by grants R01-AI14812701 from the National Institute for Allergy and Infectious Diseases to J.A.L., and R01-AI139761 from the National Institute for Allergy and Infectious Diseases to N.E.D.

