## Supplementary Material for "Use of recently vaccinated individuals to detect bias in test-negative case-control studies of COVID-19 vaccine effectiveness"

Hitchings et al

**Supplementary Methods**

*Derivation of bias-indicator*

The odds ratio comparing vaccination status between cases and controls is given by

$$OR_{U}^{R}\left( t \right)= \frac{C_{R+}\left( t \right)C_{U-}\left( t \right)}{C_{R-}\left( t \right)C_{U+}\left( t \right)}$$

$$=\frac{\pi_{-}\lambda_{-}{[\mu_{U}\alpha}_{U-}(1-v)t+\mu_{P}\alpha_{P-}vT_{V}]P}{\pi_{+}\left[ \mu_{U}\left( 1-e^{-\alpha_{U+}\lambda_{+}t} \right)\left( 1-v \right)+\mu_{P}\left( 1-e^{-\alpha_{P+}\lambda_{+}T_{V}} \right)v \right]P}\frac{\pi_{+}\mu_{R}\left( 1-e^{-\alpha_{R+}\lambda_{+}T_{P}} \right)vP}{\pi_{-}\mu_{R}\alpha_{R-}\lambda_{-}T_{P}vP}$$

$$\approx\frac{\alpha_{R+}}{\alpha_{R-}}\frac{\left[ \mu_{U}\alpha_{U-}\left( 1-v \right)t+\mu_{P}\alpha_{P-}vT_{V} \right]}{\left[ \mu_{U}\alpha_{U+}\left( 1-v \right)t+\mu_{P}\alpha_{P+}vT_{V} \right]},$$

as $1-e^{-x}\approx x$ when *x* is small.

*Derivation of bias-corrected vaccine effectiveness estimate*

The odds ratio comparing vaccination status between cases and controls is given by

$$1-OR_{R}^{F}\left( t \right)=1- \frac{C_{F+}\left( t \right)C_{R-}\left( t \right)}{C_{F-}\left( t \right)C_{R+}\left( t \right)}$$

$$= 1-\left( \frac{\pi_{+}\mu_{V}\left[ \left( 1-\varphi\right)\left( 1-e^{-\alpha_{V+}\lambda_{+}\left( t-T_{P}-T_{V} \right)} \right)+\varphi\left( 1-e^{-\alpha_{V+}\theta\lambda_{+}\left( t-T_{P}-T_{V} \right)} \right) \right]vP}{\pi_{+}\mu_{V}\left( 1-e^{-\alpha_{V+}\lambda_{+}T_{P}} \right)vP} \right)\frac{\pi_{-}\mu_{V}\alpha_{V-}\lambda_{-}T_{P}vP}{\pi_{-}\mu_{V}\alpha_{V-}\lambda_{-}\left( t-T_{P}-T_{V} \right)vP}$$

$$\approx1-\left( \frac{\left[ \left( 1-\varphi\right)\alpha_{V+}\lambda_{+}\left( t-T_{P}-T_{V} \right)+\varphi\alpha_{V+}\theta\lambda_{+}\left( t-T_{P}-T_{V} \right) \right]}{\left( \alpha_{V+}\lambda_{+}T_{P} \right)} \right)\frac{T_{P}}{\left( t-T_{P}-T_{V} \right)}$$

$$=\varphi\left( 1-\theta\right).$$

Again, $1-e^{-x}\approx x$ when x is small.

*Derivation of bias-corrected vaccine effectiveness estimate with varying T_V_*

If vaccination occurred on a set of days *T_Vj_*, and a proportion *p_j_* of vaccinated individuals is vaccinated on each vaccination day, then

$$C_{F+}\left( t \right)=\sum_{j} C_{F+}\left( t|T_{V}=T_{Vj} \right)p_{j}$$

$$C_{F-}\left( t \right)=\sum_{j} C_{F-}\left( t|T_{V}=T_{Vj} \right)p_{j}$$

Expressions for $C_{R+}\left( t \right)$ and $C_{R-}\left( t \right)$ are independent of *T_V_*, and therefore remain as in the main text. Thus,

$$1-OR_{R}^{F}\left( t \right)=1- \frac{C_{F+}\left( t \right)C_{R-}\left( t \right)}{C_{F-}\left( t \right)C_{R+}\left( t \right)}$$

$$= 1-\left( \frac{\sum_{j} [\left( 1-\varphi\right)\left( 1-e^{-\alpha_{V+}\lambda_{+}\left( t-T_{P}-T_{Vj} \right)} \right)+\varphi\left( 1-e^{-\alpha_{V+}\theta\lambda_{+}\left( t-T_{P}-T_{Vj} \right)} \right)]p_{j}}{\left( 1-e^{-\alpha_{V+}\lambda_{+}T_{P}} \right)} \right)\frac{T_{P}}{\sum_{j} (t-T_{P}-T_{Vj})p_{j}}$$

$$\approx1-\left( \frac{\alpha_{V+}\lambda_{+}[\left( 1-\varphi\right)+\varphi\theta]\sum_{j} (t-T_{P}-T_{Vj})p_{j}}{\alpha_{V+}\lambda_{+}T_{P}} \right)\frac{T_{P}}{\sum_{j} (t-T_{P}-T_{Vj})p_{j}}$$

$$=\varphi\left( 1-\theta\right).$$
